# The contribution of *RBM20* truncating variants to human cardiomyopathy

**DOI:** 10.1101/2025.07.26.25332081

**Authors:** Brendan J. Floyd, Joyce N. Njoroge, Vikki A. Krysov, Bruna Gomes, Ryan Murtha, Chiaka Aribeana, Douglas Cannie, Eric Smith, Alessia Paldino, Emily E. Brown, Andreas Barth, Erkan Ilhan, Renee Johnson, Julianne Wojciak, Mohamad Alkhayat, Sharon Graw, Kristen Medo, Jan Haas, C. Anwar A. Chahal, Kai Fenzl, Lars Steinmetz, Michael Gollob, Euan Ashley, Sharlene Day, Daniel Judge, Jason Roberts, Vasanth Vedantham, Chad Y. Mao, Diane Fatkin, Neal K. Lakdawala, Matthew R. G. Taylor, Luisa Mestroni, Ardan M. Saguner, Upasana Tayal, Julia Cadrin-Tourigny, Andrew D. Krahn, Cynthia James, Matteo Dal Ferro, Gianfranco Sinagra, Marco Merlo, Anjali Owens, Nosheen Reza, Sara Saberi, Adam Helms, Perry Elliott, Benjamin Meder, Victoria N. Parikh

## Abstract

**Background:** Genetic diagnosis has become increasingly important to guide clinical decision making for patients with dilated cardiomyopathy (DCM). Disease-causing (P/LP) missense variants in the gene *RBM20* cause a highly penetrant arrhythmogenic dilated cardiomyopathy (DCM), but the role of truncating *RBM20* variants (*RBM20tvs*) is unclear.

**Objective:** Assess the contribution of *RBM20tvs* to DCM.

**Methods:** We assembled an international cohort of DCM patients with *RBM20* variants and used data from the genome-first UK Biobank (UKB) to assess the etiologic fraction, natural history and penetrance of *RBM20tvs*.

**Results:** The etiologic fraction of *RBM20tvs* in arrhythmogenic DCM was modest (0.53[0.32,0.67], p=7.5×10^-5^). *RBM20tv* DCM patients presented to referral centers later in life than *RBM20* P/LP DCM patients (53±10 vs. 34±18 years, p=4×10^-3^), and were less likely to have a family history of sudden cardiac arrest (20% vs. 65%, p= 0.046) or cardiomyopathy (20% vs. 78% p=5.4×10^-3^). There was no significant difference in age- and sex-adjusted incident major heart failure or arrhythmia events between *RBM20tv and RBM20* P/LP DCM patients, though sex-adjusted lifetime hazard was reduced in *RBM20tv* DCM (HR 0.15[0.03,0.66],p=0.009). In UKB, lifetime incidence of cardiomyopathy, heart failure, or major ventricular arrhythmia diagnosis was lower in participants with *RBM20tvs* than in those with *TTNtvs* (HR 0.55 [0.36,0.84], p=5.9×10^-3^).

**Conclusions:** *RBM20tvs* contribute to arrhythmogenic DCM phenotypes, but confer milder disease severity alone than *RBM20* P/LP variants, and reduced lifetime disease penetrance compared to *TTNtvs*. Their potential for additive interactions with other damaging variants should be considered in DCM patients and families.

## Introduction

Within the highly morbid spectrum of systolic heart failure (HF), nonischemic dilated cardiomyopathy (DCM) accounts for 40% of diagnosis.(1,2) Familial DCM is common among these patients, and precise genetic diagnosis can guide estimation of sudden cardiac arrest (SCA) risk and prognostication of end-stage heart failure (ESHF) requiring advanced therapies.(3–8) Causative variants in several genes been associated with a risk of SCA out of proportion to the severity of reduced left ventricular ejection fraction (LVEF). Primary prevention implantable cardioverter-defibrillator (ICD) is currently recommended at earlier stages of systolic dysfunction for DCM patients with pathogenic variants in particularly arrhythmogenic genes.(9) Emerging evidence suggest that this group should be expanded to include pathogenic variants in the gene RNA binding motif 20 (*RBM20*).(10)

Disease causing missense *RBM20* variants are present in ∼3-6% of DCM cases (11–13) and are associated with a highly penetrant arrhythmogenic DCM.(14–16) Overall incidence of HF and early ventricular arrhythmias in *RBM20* DCM is high, and disease progression to ESHF appears to be sex-dependent.(10) However, the disease contribution of truncating variants in *RBM20* (*RBM20tvs*) remains unclear. The mechanism of disease-causing *RBM20* missense variants appears to be dominant negative, related to the formation of toxic ribonucleoprotein aggregates, rather than the loss of function mechanism expected with early protein truncation.(17–19) Animal models carrying *RBM20* deletions appear to have a milder phenotype than missense variant animals.(20) Further, small patient cohorts have reported later age at diagnosis for *RBM20tv* DCM patients as compared to patients with disease-causing missense variants.(21,22) The population attributable risk (etiologic fraction) of *RBM20tvs* in human DCM has yet to be estimated, and the penetrance and longitudinal impact of these variants compared to other DCM-causing variants remains poorly understood.

Here we assess the etiologic fraction of *RBM20tvs* in arrhythmogenic DCM and evaluate the natural history of *RBM20tv* DCM in an international *RBM20* DCM registry. We go on to compare the penetrance of arrhythmogenic DCM diagnoses in *RBM20tvs* to the most common genetic cause of DCM, truncating variants in titin (*TTNtvs*),(23,24) in the UK Biobank (UKB).

## Methods

### Assembly, variant curation and event definitions in International RBM20 DCM Cohort

Patients with a variant in *RBM20* identified on clinical genetic testing were identified at international familial cardiomyopathy centers: Barts Heart Centre (London, UK), Brigham and Women’s Hospital (Massachusetts, USA), Children’s Healthcare of Atlanta (Georgia, USA), University of Colorado Anschutz Medical Campus (Colorado, USA), Heidelberg University (Germany), Johns Hopkins University (Maryland, USA), University of Pennsylvania (Pennsylvania, USA), Royal Brompton Hospital (London, UK), Stanford University (California, USA), University of Trieste (Italy), University of British Columbia (Canada), Victor Chang Cardiac Research Institute (Australia), University of California, San Francisco (California, USA), Wellspan Health (Pennsylvania, USA), University of Michigan (Michigan, USA), and University of Zurich (Switzerland). Clinical data were collected retrospectively from real-world prospective care at these centers in casenote abstraction forms. Dates were not provided but recorded as ages in years. Variables included: baseline demographics, measurements from echocardiography, electrocardiogram, and cardiac magnetic resonance imaging, and clinical events occurring prior to initial evaluation and at the most recent follow up. Data were collected, stored, and shared in a deidentified manner in accordance with the institutional review board at each contributing center.

Variant pathogenicity was adjudicated centrally by a licensed clinical genetic counselor and a board certified medical geneticist. Each variant was classified into one of four categories based on American College of Medical Genetics criteria with modifications for DCM: pathogenic/likely pathogenic variants (P/LP), variants of uncertain significance (VUS), truncating variants, and benign/likely benign variants (**See Supplemental File 1)**.(25) Individuals without genetic data, or who also carried a pathogenic variant in a different established cardiomyopathy gene listed by the ClinGen expert panel on dilated cardiomyopathies were excluded.(26) Of the 193 individuals enrolled with *RBM20* variants, 14 were excluded: two of these were due to the presence of a pathogenic variant in another cardiomyopathy gene, and the remaining 12 were excluded because they carried an *RBM20* VUS with ambiguous impact on splicing which prevented further determination of whether the variant may lead to a truncated protein product. VUS with ambiguous impact on splicing were identified based on the following characteristics: within 3 nucleotides of the exon-intron junction but with low SpliceAI score (<0.05) (5 individuals), in-frame insertions or deletions (4 individuals), intronic variants with borderline SpliceAI score (0.1-0.2) (2 individuals), and one individual with a multi-exonic duplication involving the terminal exons.

Primary outcomes for survival analyses included three composite outcomes: (1) major heart failure events (MHF; defined by HF hospitalization (hospital admission with an HF-associated diagnosis), orthotopic heart transplantation (OHT), or left ventricular assist device (LVAD) implantation); (2) major ventricular arrhythmia events (MVA, defined by sustained ventricular tachycardia, appropriate ICD shock, sudden cardiac arrest or sudden cardiac death); and (3) combined major HF and MVA (MHFVA). All deaths in this cohort occurred following an MVA or HF event, and so death was not included in composite outcomes.

### Variant filtering and diagnosis definitions in clinical genetic testing cohort and gnomAD

To determine the enrichment of *RBM20tvs* in arrhythmogenic DCM (etiologic fraction), we used data from a de-identified clinical genetic testing cohort (provided via collaboration with Invitae, Inc).(27) This cohort included 314 patients with genetic variants in *RBM20*. ICD-10 codes and/or clinical indications were reviewed for diagnoses relevant to arrhythmogenic DCM to identify patients for the disease cohort as follows: personal and family history of sudden cardiac arrest/death (I46, Z82.41, Z86.74), cardiac arrhythmias including Brugada and Long QT syndrome (I49.8, I45.81), paroxysmal atrial fibrillation (I48.0), catecholaminergic polymorphic ventricular tachycardia (I47.2), supraventricular tachycardia, sustained and non-sustained ventricular tachycardia (I47.1, I47.2, I47.3), ventricular fibrillation (I49.01), nonischemic cardiomyopathies (I42.8, DCM (I42.0), hypertrophic cardiomyopathy (I42.2), left ventricular non-compaction (I42.8), peripartum (O90.3), arrhythmogenic right ventricular (I42.8), unspecified (I43)), congestive heart failure (I50), heart transplant (Z94.1), and enlarged heart (I51.7). Individuals without an arrhythmogenic DCM-relevant diagnosis, or who were missing an ICD-10 code and clinical indication, were excluded. Thus, 216 *RBM20* variant probands met criteria for an arrhythmogenic DCM diagnosis. Any patients with an additional pathogenic variant were excluded, yielding 137 probands whose data were utilized for subsequent analyses. For the general population comparator, a total of 3,978 *RBM20* variants were identified in the general population using the genome aggregation database (gnomAD v4.1.0, GRCh38). (28)

Variants (both from the clinical genetic testing cohort and in gnomAD) were filtered by variant type and variants with gnomAD minor allele frequency (MAF) >0.0001 were excluded from further analyses. After this filter, among the clinical genetic testing cohort variants, 4 were classified as truncating variants (frameshift or stop-gain). Within the general population cohort, 112 truncating (stop gain and frameshift) variants were identified. Splice variants from both groups were excluded given uncertainty around their heterogeneity of effect on truncation.

### Variant filtering and diagnosis definitions in UK Biobank

*RBM20* and *TTN* variants with gnomAD v3 minor allele frequency <0.005 were identified among the UKB cohort of 502,132 participants with genome data using the UK Biobank Browser.(4,29) For *RBM20*, all frameshift, nonsense, and synonymous variants were identified. The negative control population was defined as carriers of *RBM20* synonymous variants (excluding those with any SpliceAI score above 0). *TTNtvs* were identified as frameshift and nonsense variants within the A-band (GRCh38 2:178,535,982-178,618,491). Splice variants were excluded from both *RBM20* and *TTN* for this analysis given uncertainty regarding their heterogeneous effect. Genome variant files were obtained covering the exons and exon-intron junctions for *RBM20* and *TTN*. In total, 174 RBM20tvs, 965 *TTNtvs*, and 3110 *RBM20* synonymous variant carriers were identified (**Supplemental Table 1**).

Demographic and clinical data, including sex, birth year, birth month, age at enrollment, date of death, and dates of first reported diagnoses of cardiomyopathy (ICD10 code I42), DCM (I42.0), cardiac arrest (I46), heart failure (I50), atrial fibrillation and flutter (I48), other cardiac arrhythmias (I49), obesity (E66), hypertension (I10-I13 and I15), type 2 diabetes mellitus (E10-E14), and coronary artery disease (Z95.1, Z95.5, R93.1) were recorded. Three composite outcomes were evaluated: cardiomyopathy or heart failure diagnosis (codes I42, or I50), arrhythmia diagnosis (codes I46, or I49), and arrhythmogenic DCM diagnosis (codes I42, I46, I49, and I50). Atrial fibrillation and atrial flutter (I48) diagnoses were evaluated in a separate analysis.

### Statistical Comparisons

Etiologic fractions were calculated as a function of the odds ratio: (odds ratio - 1)/odds ratio, to determine the attributable risk of RBM20tvs to arrhythmogenic DCM (defined as above) or DCM phenotype (defined as ICD 10 code I42.0 specifically).(27) For data collected at initial visit with a familial DCM center and baseline characteristics in UKB, means were compared using a student’s T-test and proportions were compared using Chi-squared or Fisher’s exact test (for N<100). Time-to-event data from the International *RBM20* DCM Cohort and in the UKB were compared at last follow-up using multivariate Cox regression analyses adjusting for age and/or sex as indicated in results. In all comparisons, patients with missing values for that comparison were excluded.

## Results

### *RBM20tvs* account for small etiologic fraction of arrhythmogenic DCM

To assess the association between *RBM20tvs* and arrhythmogenic DCM we determined the etiologic fraction (EF) of *RBM20tvs* associated with arrhythmogenic DCM phenotypes in a clinical genetic testing cohort with limited phenotyping (diagnostic codes, N=4) vs. the general population (gnomAD v4.1.0, N=327).(28) This small number of *RBM20tvs* was not significantly associated with arrhythmogenic DCM diagnoses (OR 3.0 [0.8-7.8], EF 0.67 [0-0.87], p=0.05). Given limited sample size, we sought to replicate our results in the UK Biobank (UKB). We identified 174 carriers of truncating variants in *RBM20* in UKB. Among the carriers of RBM20tvs, 41 carried an ICD-10 diagnosis associated with arrhythmogenic DCM (24%). RBM20tvs were mildly enriched in participants with arrhythmogenic DCM-relevant diagnoses as compared to participants without arrhythmogenic DCM-relevant diagnoses (OR 2.1 [1.5-3.1], EF 0.53 [0.32-0.67], p=7.5×10^-5^) **(Table 1).** We repeated this analysis restricting only to the ICD-10 code for DCM (I42.0), and found that 1.8% of RBM20tv carriers had this diagnosis compared to 0.32% of the UKB at large, demonstrating a greater enrichment for this more specific diagnosis (OR 5.59 [1.78-17.5], EF 0.82 [0.44-0.94], p= 0.003). *TTN* truncating variants (TTNtvs) were enriched more than RBM20tvs both for composite arrhythmogenic DCM diagnoses (OR 3.5 [3.1-4.0], EF 0.71 [0.67-0.75], p<1×10^-7^) and for DCM-specifically (OR 54.0 [42.1-69.2], EF 0.98 [0.95-0.98], p<1×10^-7^). *RBM20* synonymous variants were not enriched in participants with an arrhythmogenic DCM diagnosis (OR 1.0 [0.91-1.1],p=0.77).

**Table 1:** Etiologic fraction of RBM20tvs for arrhythmogenic DCM diagnoses. ^a^ Odds ratio and p-value vs gnomAD v4.1.0. ^b^ Odds ratio and p-value vs the entire UK Biobank cohort. ^c^ Fisher’s exact test p-value. Key: CI: confidence interval; EF: etiologic fraction, UKB: UK Biobank, NS: not significant, p-value >0.05.

| Variant type | Cohort | Odds ratio | Etiologic fraction | p-value <sup>c</sup> |
| --- | --- | --- | --- | --- |
| **RBM20tv** | Clinical genetic testing<sup>a</sup> | 3.00<br>(0.81-7.77) | 0.67<br>(0-0.87) | 0.05 |
| **RBM20tv** | UKB <sup>b</sup> | 2.15<br>(1.48 - 3.08) | 0.53 (0.32-0.67) | 7.45 x10<sup>-5</sup> |
| ***RBM20***<br>**synonymous** | UKB <sup>b</sup> | 1.02<br>(0.91-1.13) | 0.02 | 0.77 |
| **TTNtv** | UKB <sup>b</sup> | 3.48<br>(3.06 - 3.98) | 0.71<br>(0.67 - 0.75) | <1x10<sup>-7</sup> |

### *RBM20*tv transcript position is not correlated with disease association

As the RBM20 arginine-serine rich (RS) domain is important for nuclear localization of the protein, and as mislocalization is one proposed mechanism of pathogenicity,(17) we next evaluated whether truncating variants C-terminal to the RS domain would have a decreased risk of arrhythmogenic DCM compared to those N-terminal to the RS domain. The distribution of arrhythmogenic DCM-related truncating variants across the *RBM20* transcript was similar to that of non-arrhythmogenic DCM-related variants **(Figure 1)**. Nearly half (N=19, 46%) of arrhythmogenic DCM-related variants clustered in the region between the N-terminus and RS domain, similar to the 50% (N=64) of non-arrhythmogenic-DCM-related variants found in the same region, indicating that the length of the truncated transcript is irrelevant to disease association, and supporting the hypothesis that these truncating variants undergo nonsense mediated decay, which results in loss of function (*χ*2(1)=0.052, p=0.82).

**Figure 1.**
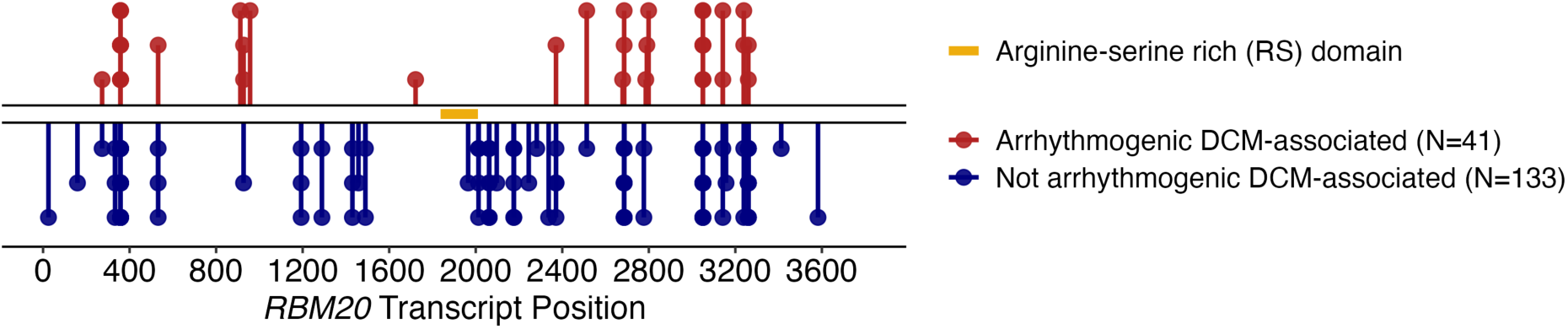
Disease-associated *RBM20tvs* are not differentially spatially distributed. Among the 174 *RBM20* truncating variant carriers identified in the UK Biobank, 41 met criteria for diagnosis of arrhythmogenic dilated cardiomyopathy (DCM) (24%) and 133 variants were not associated with arrhythmogenic DCM. Arrhythmogenic DCM-associated *RBM20* truncating variants (red) are distributed across the *RBM20* transcript similar to that of controls (blue). In relation to the arginine-serine rich (RS) domain (positions c.1839-2019, gold), nearly half (N=19, 46%) of arrhythmogenic DCM-related variants clustered in the region between the N-terminus and RS domain, comparable to the 50% (N=64) of the not arrhythmogenic DCM-associated variants found in the same region (χ^2^(1)=0.052, p=0.82).

### RBM20tv DCM probands present later and have less cardiac family history than DCM probands with disease causing missense variants in *RBM20*

Of the 179 individuals included in the International *RBM20* DCM Cohort, 50 had missense pathogenic or likely pathogenic variants (P/LP, disease causing), 96 had variants of uncertain significance (VUS), 12 had RBM20tvs, and 21 had benign or likely benign variants **(Supplemental Table 2)**. RBM20tv probands presented later in life than *RBM20* P/LP probands (RBM20tvs: 53±10 years, *RBM20* P/LP: 34±18 years, p=4.0×10^-3^) and *RBM20* VUS (*RBM20* VUS: 42±18 years, p=0.03). *RBM20tv* probands also displayed a reduced frequency of family history of SCA (20%) or DCM (20%) compared to *RBM20* P/LP probands (SCA:65% p= 0.046, DCM:78% p=5×10^-3^). Mean QRS length was also increased in *RBM20tv* patients, which has not been found to be increased in *RBM20* P/LP patients prior (p=5×10^-3^, **Table 2**)).(10,14) Clinical characteristics were otherwise similar between *RBM20tvs* and *RBM20* P/LP patients at presentation.

**Table 2:** Baseline demographics of the *RBM20* international registry. ESHF: End Stage Heart Failure; FH: Family History; HF: Heart Failure; LA: Left Atrium; LBBB: Left Bundle Branch Block LVEF: Left Ventricular Ejection Fraction; LVIDD: Left Ventricular Internal Diameter at End Diastole; MHFVA: Major Heart failure and Ventricular Arrhythmia Events; MHF: Major Heart Failure Events; MVA: Major Ventricular Arrhythmia; RBM20tv: *RBM20* truncating variant; SCA/D: Sudden Cardiac Arrest/Death; VUS: Variant of Uncertain Significance.

|  |  | Pathogenic missense | RBM20tv | Pathogenic vs RBM20tv (p-value) |
| --- | --- | --- | --- | --- |
| Individuals enrolled, N |  | 50 | 12 |  |
| Male, N (%) |  | 26 (57) | 8 (67) | 0.74 |
| Probands, N (%) |  | 20 (40) | 10 (83) | 0.01 |
| Age at presentation (years, mean(SD)) | All, mean (SD) | 31 (17) | 48 (16) | 0.002 |
|  | Probands only | 34 (18) | 53 (9.9) | 0.004 |
| FH SCA/D in probands, N (%) |  | 11 (65) | 2 (20) | 0.046 |
| FH HF in probands, N (%) |  | 14 (78) | 2 (20) | 0.005 |
| LVIDD (mm, mean, SD) |  | 51 | 57 | 0.22 |
| LVIDD indexed (mm/m<sup>2</sup>, to BSA) |  | 27 | 29 | 0.58 |
| LA volume (mm/m<sup>2</sup>, indexed to BSA) |  | 23 (6.5) | 40 (6.7) | 0.001 |
| LVEF (% , mean(SD)) |  | 42 (16) | 41 (19) | 0.77 |
| QRS (ms, mean(SD)) |  | 94 (13) | 119 (29) | 0.005 |
| QTc (ms, mean(SD)) |  | 416 (31.5) | 430 (35.7) | 0.28 |
| LBBB, n (%) |  | 0 (0) | 3 (25) | 0.0007 |
| HF Hospitalization, n (%) |  | 7 (16) | 1 (8) | 0.38 |
| Prior ESHF, n (%) |  | 2 (4) | 0 (0) | 0.32 |
| Prior MHF, n (%) |  | 9 (18) | 1 (8) | 0.43 |
| Prior MVA, n (%) |  | 3 (7) | 2 (17) | 0.54 |

DCM patients with *RBM20tvs* and *RBM20* P/LP variants had 5.0±4.8 and 7.3±6.6 years of follow up, respectively (mean±SD). DCM patients with *RBM20* VUS had 5.2±5.4 years follow up. Age- and sex-adjusted comparison of incident composite MHFVA events did not reveal a statistically significant difference between *RBM20tv* and *RBM20* P/LP DCM patients, but did reveal a lower risk in *RBM20* VUS DCM patients vs. *RBM20* P/LP DCM (*RBM20tv:* 0.37 [0.04,3.2] p=0.37; *RBM20* VUS:0.33 [0.12,0.97], p=0.04, **Supplemental Figure 1A**). Sex-adjusted comparison of lifetime composite MHFVA events displayed a significant reduction for *RBM20tv* but not *RBM20* VUS DCM patients compared to *RBM20* P/LP DCM patients (*RBM20tv:* 0.15 [0.03,0.66] p=0.01; *RBM20* VUS: 0.82 [0.48,1.43], p=0.5, **Figure 2A**). This difference was driven by a statistically significant reduction in lifetime risk of major HF events (*RBM20tv:* 0.10[0.01,0.75],p=0.03, *RBM20* VUS: 0.84[0.45,1.6],p=0.6, **Figure 2B**). No statistically significant difference was identified in rare lifelong events of major ventricular arrhythmias (*RBM20tv:* 0.17[0.02,1.33],p=0.09, *RBM20* VUS: 0.53[0.24,1.16],p=0.11, **Figure 2C).**

**Figure 2.**
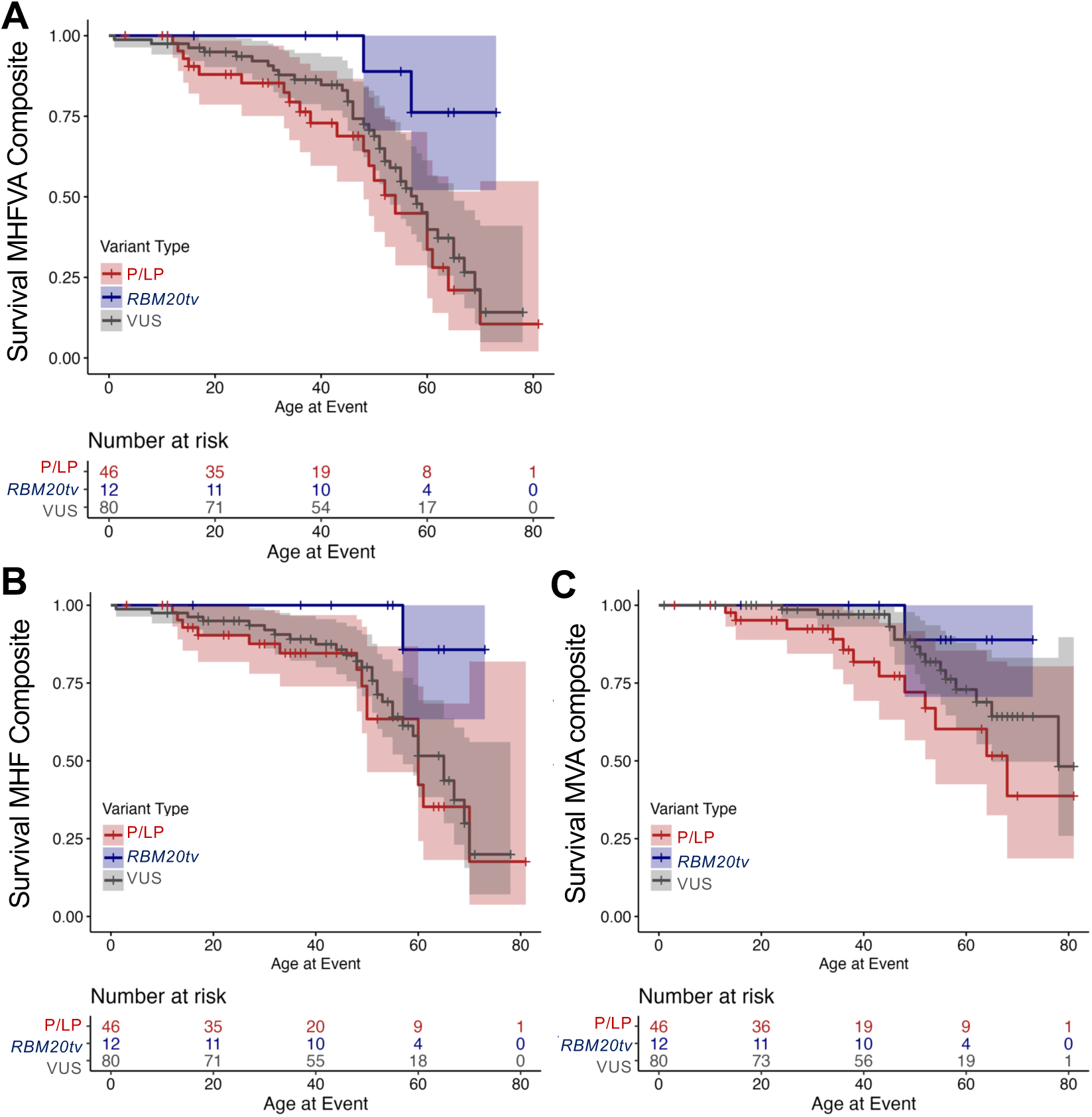
Lifetime risk of major events by *RBM20* variant type. **(A)** Risk of lifetime combined major heart failure and ventricular arrhythmia (MHFVA) events is reduced in *RBM20tv* but not *RBM20* VUS compared to disease-causing (P/LP) *RBM20* variants (HR[95%CI], *RBM20tv:* 0.15[0.03,0.66], p=0.01; *RBM20* VUS: 0.82[0.48,1.43], p=0.5.). **(B)** Risk of lifetime major heart failure (MHF) events is reduced in *RBM20tv* but not *RBM20* VUS compared to disease-causing (P/LP) *RBM20* variants (*RBM20tv:* 0.10[0.01,0.75], p=0.03, *RBM20* VUS: 0.84[0.45,1.6], p=0.6). **(C)** Risk of lifetime major ventricular arrhythmia (MVA) events does not vary significantly by variant type. (*RBM20tv:*0.17[0.02,1.33], p=0.09, *RBM20* VUS:0.53[0.24,1.16], p=0.11). HR= hazard ratio, CI= confidence interval. All analyses adjusted for sex.

### RBM20tvs show lower lifetime hazard of arrhythmogenic DCM diagnoses than TTNtv in UKB

We sought to expand our analysis to lifetime risk of arrhythmogenic DCM diagnosis in a genome-first population. No *RBM20* P/LP variants are present in UKB. We therefore examined participants in UKB with *RBM20*tvs (N=174) and compared these with participants carrying *RBM20* synonymous variants (N=3110) and those with disease causing *TTN* truncating variants (TTNtvs, N=965) (**Supplemental File 2**). There were no significant differences in age at enrollment, history of hypertension, coronary artery disease, obesity or duration of follow up across these groups. Type 2 diabetes was marginally more prevalent in the TTNtv population (**Supplemental Table 1**). The arrhythmogenic DCM composite diagnosis was experienced in 12.5% of all UKB participants (62,866 of 502,132 individuals). TTNtv carriers had an increased prevalence of this composite diagnosis compared to all UKB participants (33.3%, OR 3.48 [3.06-3.98], p-value <1×10^-5^), whereas carriers of *RBM20* synonymous variants did not (12.7%, OR 1.02 [0.91-1.13], p-value 0.77). *RBM20tv* carriers experienced a mildly increased lifetime prevalence of disease, with the composite outcome present in 23.5% of individuals (OR 2.15 [1.48-3.08], p-value <1×10^-4^). *RBM20tv* carriers experienced an increased lifetime hazard of the composite diagnosis compared to *RBM20* synonymous variant carriers (1.8 [1.2-2.8], p=0.007) but less than *TTNtv* carriers (0.5 [0.3-0.7], p=7.7×10^-4^) (**Figure 3**). This finding persisted for heart failure and cardiomyopathy diagnoses alone (vs. synonymous variants: 2.14 [1.31-3.49], p=2.39 x 10^-3^; vs. *TTNtvs*: 0.46 [0.28-0.74], p=1.56 ×10^-3^) but was not significant for arrhythmias (**Supplemental Figure 2**). A separate analysis of life-long incidence of atrial fibrillation or flutter shows *RBM20tv* carriers experienced an increased hazard compared to negative controls (1.9[1.3-2.7], p=0.001), and lower hazard than *TTNtv* carriers (0.6[0.4,0.9], p=0.02) (**Supplemental Figure 3)**.

**Figure 3.**
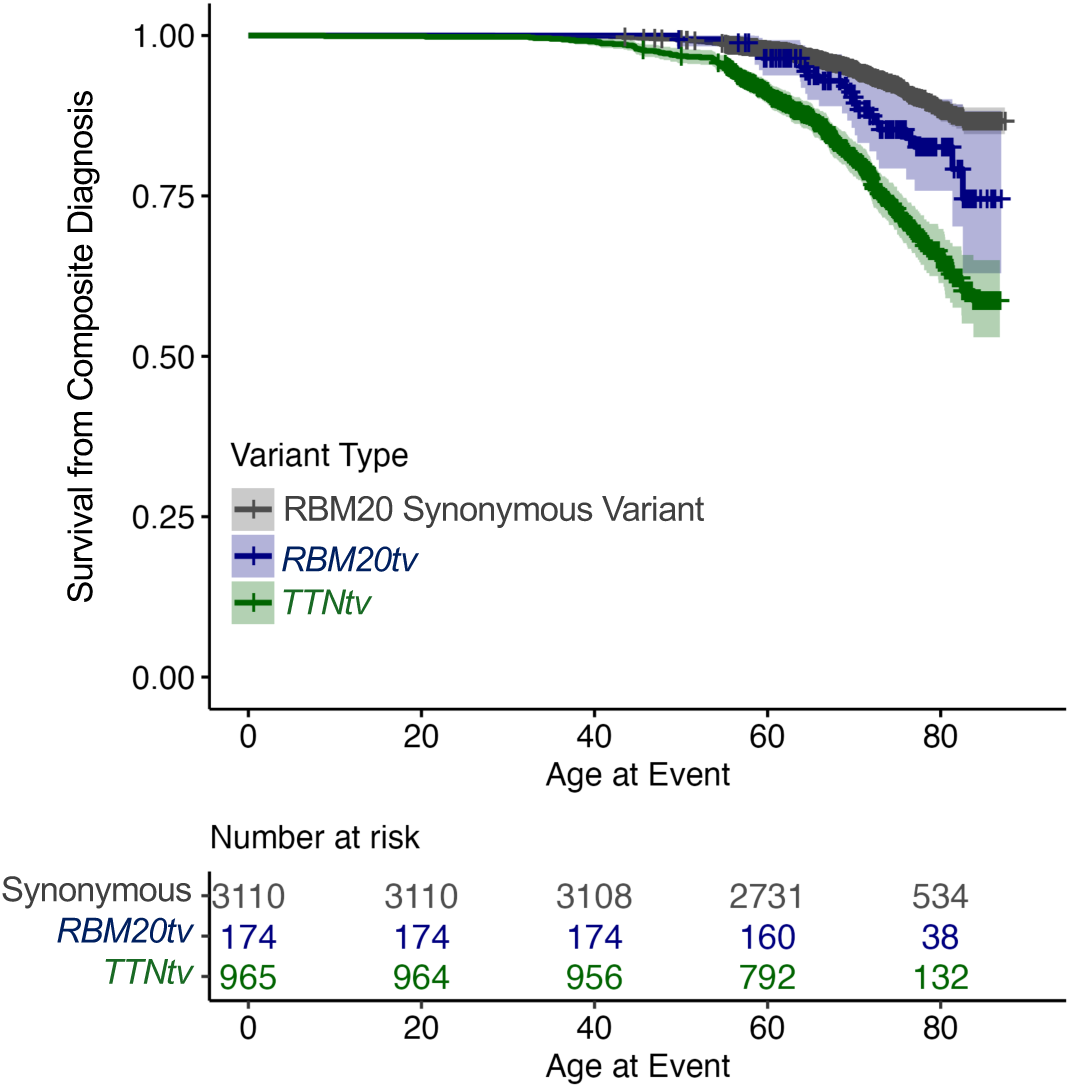
Lifetime risk of composite arrhythmogenic DCM diagnosis by variant type. *RBM20tv* carriers experienced an increased lifetime hazard of the composite diagnosis compared to *RBM20* synonymous variant carriers ( HR [95% CI] 1.8 [1.2-2.8], p=0.007) but less than *TTNtv* carriers (0.5 [0.3-0.7], p=7.7×10^-4^). Analysis adjusted for sex. HR: hazard ratio, CI: confidence interval.

## Discussion

We demonstrate here a small etiologic contribution of RBM20tvs to arrhythmogenic DCM phenotypes, reduced penetrance of *RBM20tvs* in a DCM patient population (measured by age of onset and prevalence of family history of DCM and SCA), and, lastly, reduced lifetime penetrance of heart failure in *RBM20tv* carriers with DCM in a genome-first cohort. These findings, bolstered by prior small baseline studies of *RBM20tv* DCM patients, answer critical clinical questions about the role of *RBM20tvs* in human cardiomyopathy.(21,22) They indicate that it is unlikely that *RBM20tvs* have a large independent effect on the penetrance and natural history human arrhythmogenic DCM; that is, they likely represent minor contributors to the development of arrhythmogenic DCM in the population. In DCM patients with an *RBM20tv*, they indicate the potential for a less severe disease course than P/LP missense variants in *RBM20* driven mainly by reduced major HF events. They do not at this time support the inclusion of *RBM20tvs* as a risk factor for sudden cardiac death in DCM, though larger longitudinal cohorts are needed to definitively answer that question.

As we move toward a genome-first future and expand genetic testing to more and more patients with DCM, understanding the disease contributions of classes of variants like *RBM20tvs* is critical. We find, for example, that *RBM20tvs* are less associated with life-long HF, MVA and atrial fibrillation/flutter diagnosis than*TTNtvs. TTNtvs*, while the most common cause of genetic DCM, have not been associated in large cohorts with as severe outcomes as, for example, highly arrhythmogenic and penetrant Laminopathies (LMNA).(24,30–32) Here we find that RBM20tvs have even less severe disease course than TTNtvs (though more events than a negative control cohort with synonymous variants in *RBM20*). It is notable that a larger fraction of DCM specifically is attributable to RBM20tvs than the more diffuse phenotypes included in the arrhythmogenic DCM diagnosis here. Combined with outcome differences driven by HF in UKB, this may indicate that *RBM20tvs* are also less arrhythmogenic than their missense P/LP counterparts, though larger numbers of RBM20tv DCM patients are needed to make a robust comparison. Combined with the small etiologic fraction of arrhythmogenic DCM and DCM alone explained by *RBM20tvs*, we would suggest that *RBM20tvs* be classified as low-effect contributors to disease (i.e., disease modifiers).

Why ostensible loss of function (truncating) variants in *RBM20* would have smaller etiologic contribution and milder disease course than disease causing missense variants in the RS domain and E-rich domain is an important question. While the mechanism of action of missense variants in the RS domain of *RBM20* has been reported to stem from nuclear mislocalization from the RBM20 protein, there is also significant evidence that it may contribute in a dominant negative fashion to disease, potentially via the formation of cytotoxic granules.(17,33,34) The explanation for why *RBM20tvs* would have less effect than reported variants in the E-rich domain is less straightforward. Report of a single, well-validated causative variant in this region (p.E913K), points to reduced RBM20 protein expression as its disease mechanism.(35) Regardless, our findings are in line with prior animal studies: *RBM20* knockout rodents (heterozygous and homozygous) also show relatively mild cardiac dysfunction compared to known pathogenic missense *RBM20* variants. (16,36). While the mechanism of action of many arrhythmogenic DCM genes is reduced production of functional protein, most of these are structural proteins, without an abundance of which the cardiomyocyte cannot function. The baseline expression of *RBM20* at the level of both RNA and protein in cardiomyocytes is quite low relative to these large structural proteins, and may be more amenable to stabilization of protein expression from a single functional copy. It is also possible that during stress, cardiomyocytes need to upregulate *RBM20* expression to compensate, and that having a single functional copy of *RBM20* due to an *RBM20tv* confers risk only in the setting of such a ‘second hit’.

The findings of this study exist in the context of its necessary limitations. We have attempted to balance the bias introduced by studying patients referred to tertiary care centers by also evaluating our hypotheses in a genome-first cohort (UKB), which overall selects for less morbidity than disease cohorts. We are unable, therefore, to assess the disease-modification of other genetic DCMs by RBM20tvs, which remain, though more common than *RBM20* P/LP variants, rare in the population. The absolute estimates of etiologic fraction we make using diagnosis codes in UK Biobank may ultimately slightly underestimate the effect size, as diagnostic codes are a relatively blunt diagnostic instrument, however, using the same method to calculate the etiologic fraction of TTNtvs, we show that RBM20tvs have a comparatively lower attributable risk. Our sample of DCM patients with either RBM20tvs or *RBM20* P/LP variants was also quite size-limited despite the large number of participant institutions, underlining the difficulty of studying rare disease, and the need for focused collaborative efforts to bring together larger cohorts for future study.

These data represent a significant step in understanding the disease contribution of *RBM20tvs* to arrhythmogenic DCM and interpreting their impact in the cardiovascular genetics clinic. Importantly, within the international patient cohort we present here, unlike HF events, MVA did occur before age 60 in RBM20tv patients. Even for *RBM20* P/LP variants, risk factors for MVAs remain unknown beyond mild-moderate reductions in left ventricular ejection fraction.(10) Even larger international efforts are needed to understand the risk structure of ventricular arrhythmias in *RBM20* P/LP variants and whether RBM20tvs confer additive arrhythmogenic risk to other causes of DCM.

## Data Availability

All data produced in the present study are available upon reasonable request to the authors

## Clinical Perspectives

DCM patients with *RBM20tvs* have a milder disease course than those with P/LP variants in RBM20, and display lower life-time penetrance of arrhythmogenic DCM-related diagnoses than *TTNtvs* in a genome-first population.

## Conflicts of Interest/Disclosures

VP is an SAB member of Lexeo Therapeutics and SolidBio and has minor consulting or advisory relationships with Borrealis, Constantiam Biosciences, and BioMarin, from which she also receives research sponsorship. NL receives research support from BMS and Pfizer and modest consulting incomes from BMS, Pfizer, Alexion, Nuevocore, Tenaya, Cytokinetics, and Gemma. AMS received speaker /advisory board /consulting fees from Medtronic and Zoll. LM is consultant for Tenaya, Alexion, Rocket Pharma, and received research support from Greenstone Bio, Pfizer. UT is a consultant (research editor) for The BMJ. AK is a Consultant to Tenaya Therapeutics. CAJ receives research funding from Lexeo Therapeutics, Rocket Pharmaceuticals, ARVADA Therapeutics, and Tenaya Therapeutics.

## Funding and Acknowledgements

First we want to thank our patients for generously participating in research. This work was supported by the National Institutes of Health R01HL168059 (VP), R01HL164675 (VP and EA), R01HL164634, R01HL147064, R01HL170012 (LM and MRGT) and Foundation LeDucq Transatlantic Network of Excellence (VP, EA and LS). The Zurich ARVC Program (AS) is supported by the Georg und Bertha Schwyzer-Winiker Foundation, Baugarten Foundation, USZ Foundation (Dr. Wild Grant), Swiss Heart Foundation (grant no. FF17019 and FF21073 to AMS), and Swiss National Science Foundation (grant No. 160327 to FD and 10.001.787 to AMS).

## SUPPLEMENTAL INFORMATION

### Supplemental tables

**Supplemental Table 1:** Demographics of UK Biobank cohort. ^a^ Fisher’s exact, RBM20tv vs *RBM20* synonymous, ^b^ One-way ANOVA ^c^ ICD10 codes used: Z95.1, Z95.5, or R93.1. Abbreviations: RBM20tv:*RBM20* truncating variant; SD: Standard Deviation; TTNtv: *TTN* truncating variant

|  | RBM20tv | *RBM20*<br>synonymous | TTNtv | p-value |
| --- | --- | --- | --- | --- |
| Age at recruitment, y<br>(mean, SD) | 57.3 (7.8) | 56.1 (8.3) | 56.2 (8.1) | 0.16 <sup>b</sup> |
| Age at death or last<br>follow up, y (mean, SD) | 73.0 (8.1) | 72.0 (8.2) | 72.1 (8.0) | 0.27 <sup>b</sup> |
| Time followed, y<br>(mean, SD) | 15.7 (2.5) | 15.9 (2.4) | 15.8 (2.6) | 0.44 <sup>b</sup> |
| Deceased, N (%) | 19 (11) | 256 (8) | 109 (11) | 0.21 <sup>a</sup> |
| Age at death, y (mean,<br>SD) | 68.3 (7.7) | 71.2 (8.1) | 72.2 (6.8) | 0.10 <sup>b</sup> |
| Hypertension, N (%)<br>ICD10 codes I10-I13<br>and I15 | 69<br>(38.2) | 1254<br>(40.3) | 271<br>(51.6) | 0.87 <sup>a</sup> |
| Obesity, N (%)<br>ICD10 code E66 | 23<br>(11.7) | 326<br>(10.5) | 84<br>(16) | 0.26 <sup>a</sup> |
| Diabetes mellitus, N<br>(%)<br>ICD10 codes E10-E14 | 12<br>(7.1) | 388<br>(12.5) | 108<br>(20.6) | 0.04 <sup>a</sup> |
| History of coronary<br>artery disease, N (%) <sup>c</sup> | 9<br>(5.2) | 118<br>(3.8) | 52<br>(9.9) | 0.32 <sup>a</sup> |

**Supplemental Table 2:**
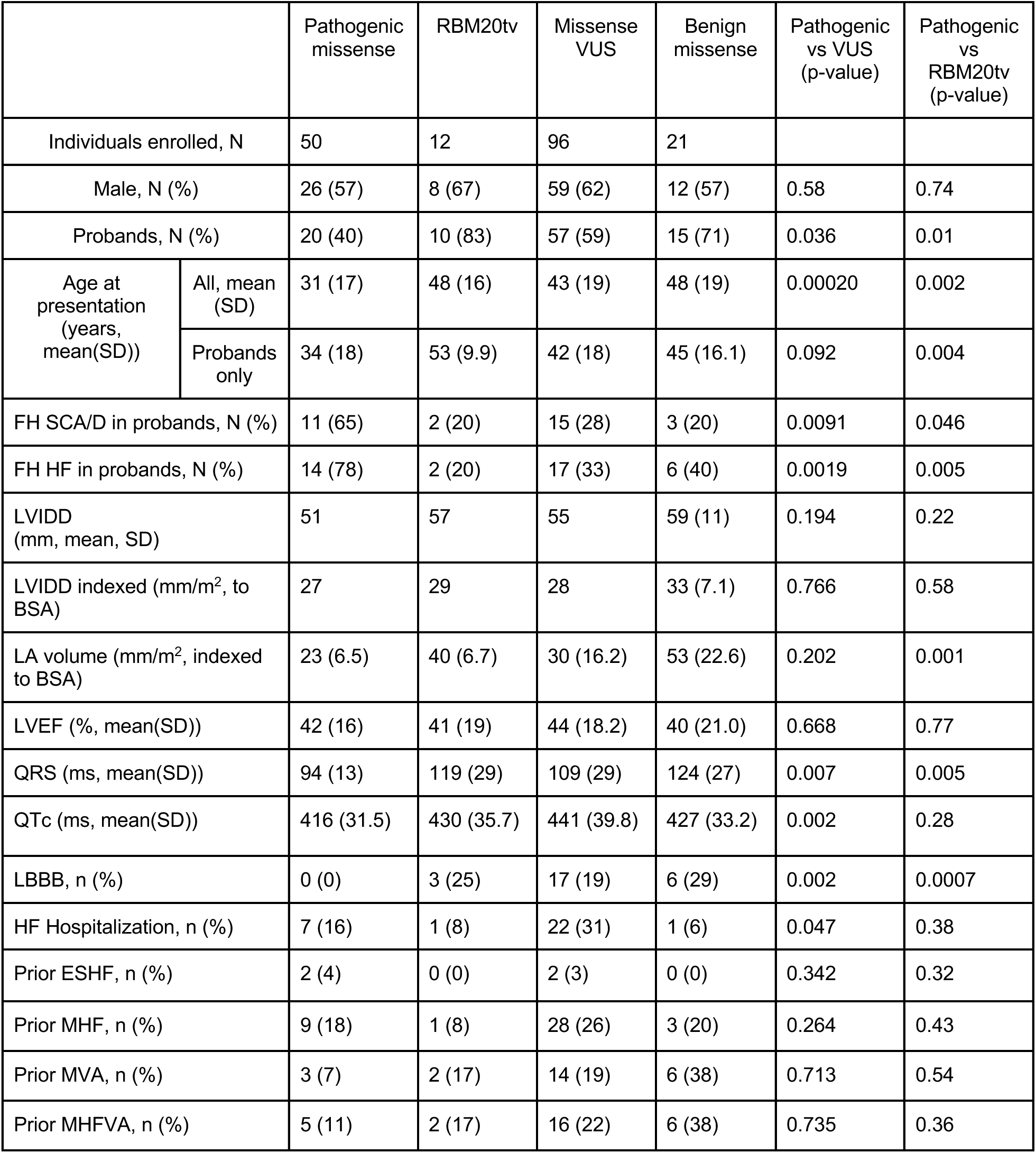
Baseline demographics of the *RBM20* international registry. Abbreviations: ESHF: End Stage Heart Failure; FH: Family History; HF: Heart Failure; LA: Left Atrium; LBBB: Left Bundle Branch Block LVEF: Left Ventricular Ejection Fraction; LVIDD: Left Ventricular Internal Diameter at End Diastole; MAHE: Major adverse Arrhythmic and Heart failure Events; MHFE: Major Heart Failure Event; MVA: Major Ventricular Arrhythmia; RBM20tv: *RBM20* truncating variant; SCA/D: Sudden Cardiac Arrest/Death; VUS: Variant of Uncertain Significance

### Supplemental Figures

**Supplemental Figure 1.**
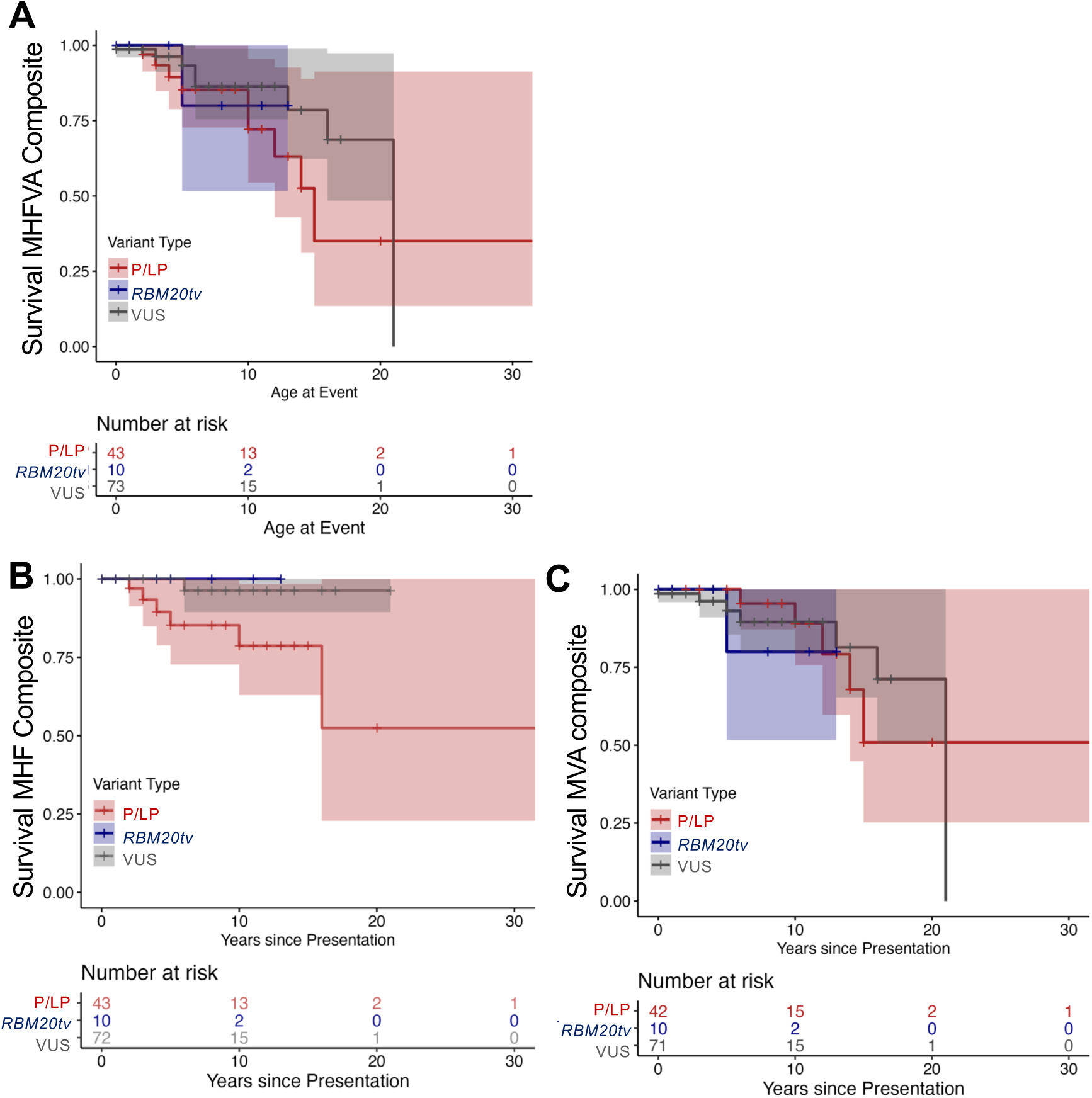
Incident major events by *RBM20* variant type. **(A)** Risk of combined major heart failure and ventricular arrhythmia (MHFVA) events is reduced in *RBM20* VUS compared to disease-causing (P/LP) *RBM20* variants but no statistically significant difference is found for *RBM20tv* (HR[95%CI], *RBM20* VUS: 0.33 [0.12,0.96], p=0.04; *RBM20tv:* 0.37 [0.04,3.2], p=0.37). **(B)** Risk of incident major heart failure (MHF) events is reduced in *RBM20* compared to disease-causing (P/LP) *RBM20* variants (*RBM20* VUS: 0.08 [0.01,0.69], p=0.02). No incident MHF events occurred in *RBM20tv* patients. **(C)** Risk of incident major ventricular arrhythmia (MVA) events does not vary significantly by variant type. (*RBM20tv:*1.1[0.11,11.1], p=0.93, *RBM20* VUS:0.70[0.2,2.5], p=0.59). HR= hazard ratio, CI= confidence interval. All analyses adjusted for age and sex.

**Supplemental Figure 2.**
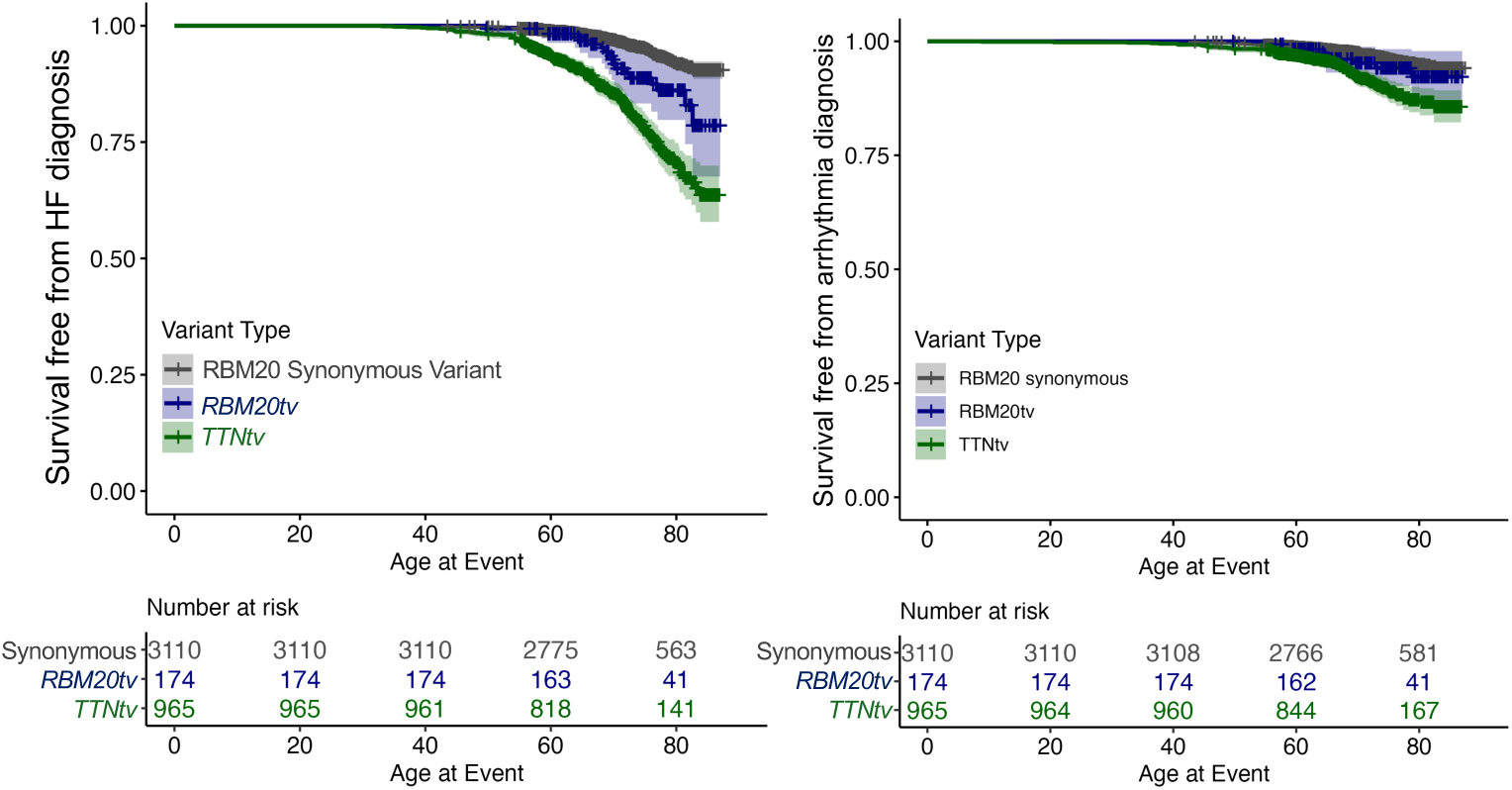
Lifetime composite HF/cardiomyopathy and arrhythmia diagnosis stratified by variant type. **(Left)** Risk of heart failure or cardiomyopathy (HF) diagnoses is higher in carriers of RBM20tv vs *RBM20* synonymous variants (HR [95% CI] 2.14 [1.31-3.49], p=2.39 x 10^-3^) but lower than in TTNtv carriers (HR 0.46 [0.28-0.74], p=1.56 ×10^-3^). **(Right)** Risk of major arrhythmias is not significantly different in carriers of *RBM20tv* vs *RBM20* synonymous variants (HR [95% CI] 1.50 [0.76-2.96], p=0.25) and lower than in *TTNtv* carriers (HR 0.65 [0.45-0.93], p=0.0175). HR= hazard ratio, CI= confidence interval. Analysis adjusted for sex.

**Supplemental Figure 3:**
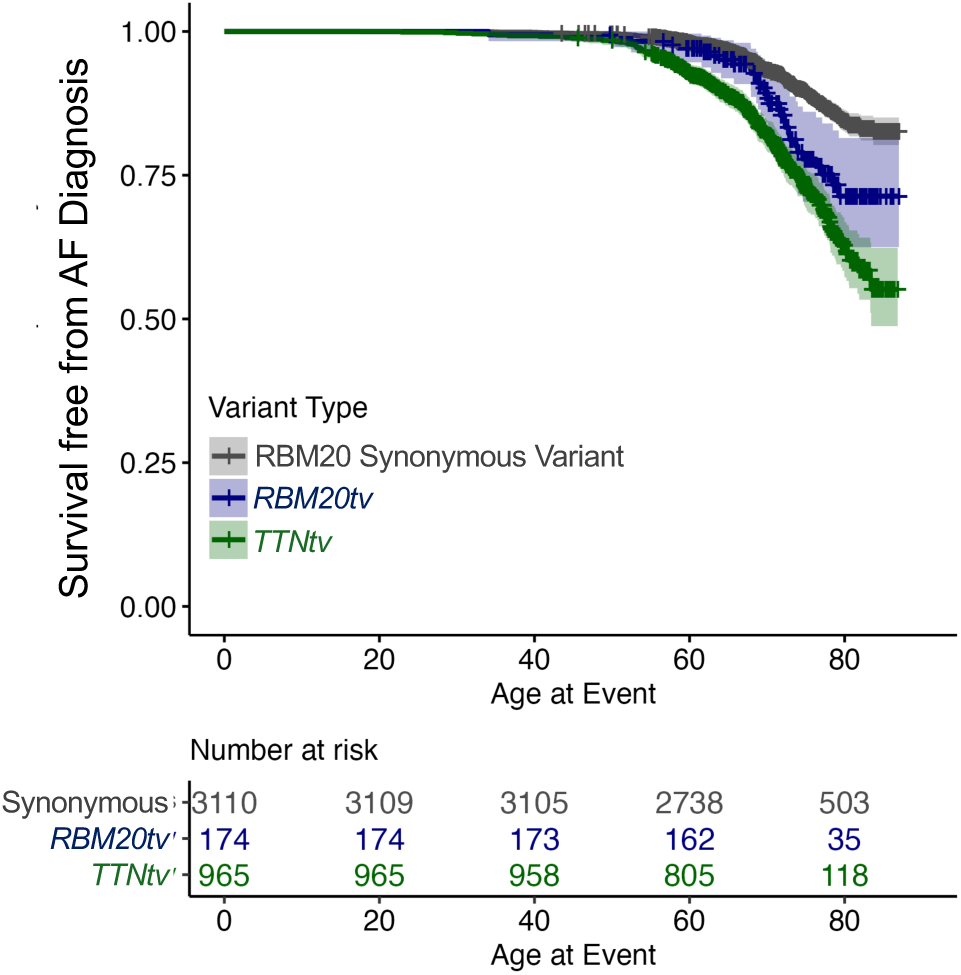
Lifetime atrial fibrillation or flutter (AF) diagnosis stratified by *RBM20* and *TTN* variant type. Risk of atrial fibrillation or flutter diagnosis is higher in carriers of *RBM20tv* vs *RBM20* synonymous variants (HR [95% CI] 1.9 [1.3-2.7], p=0.001) and lower than *TTNtv* carriers (0.6 [0.4-0.9], p=0.02). HR: hazard ratio, CI: confidence interval. Analysis adjusted for sex.

## Notes

### Author Declarations

The IRB of Stanford University gave ethical approval for this work

